# The small molecule inhibitor 3PO is a modulator of neutrophil metabolism, ROS production and NET release

**DOI:** 10.1101/2023.12.02.23299318

**Authors:** Michele Fresneda Alarcon, Genna Ali Abdullah, Andy Nolan, Christina Linford, Maria Martina Meschis, Andrew L Cross, Andrew Sellin, Marie M Phelan, Helen L Wright

**Author notes:** Correspondence: Dr Helen L Wright.

## Abstract

Neutrophils are key effector leukocytes of the innate immune system and play a pivotal role in defending the host against microbial infections. Recent studies have identified a crucial link between glycolysis and neutrophil cellular functions. Using human neutrophils, we have investigated the intricate relationship between glycolysis, extracellular glucose availability, and the enzyme 6-phosphofructo-2-kinase/fructose-2,6-bisphosphatase 3 (PFKFB3), in the regulation of reactive oxygen species (ROS) and neutrophil extracellular trap (NET) production. We have identified that PFKFB3 is elevated in rheumatoid arthritis (RA) neutrophils and that the small molecule PFKFB3 inhibitor 3PO is a key regulator of neutrophil ROS and NET production. 3PO blocked the production of ROS and NETs in a dose-dependent manner in both RA and healthy (HC) neutrophils (p<0.01), and RA neutrophils were more sensitive to lower concentrations of 3PO. Bacterial killing was only partially inhibited by 3PO, and the proportion of live neutrophils after 24h incubation was unchanged. Using NMR metabolomics, we identified that 3PO increases the concentration of lactate, phenylalanine and L-glutamine in neutrophils, as well as significantly decreasing intracellular glutathione (adj. p-value<0.05). We also demonstrated that RA neutrophils produce ROS and NETs in culture conditions which mimic the low glucose environments encountered in RA synovial joints. Our results also suggest 3PO may have molecular targets beyond PFKFB3. By dissecting the intricate interplay between metabolism and neutrophil effector functions, this study advances the understanding of the molecular mechanisms governing pro-inflammatory neutrophil responses and identifies 3PO as a potential therapeutic for conditions characterized by dysregulated neutrophil activation.

## 2. INTRODUCTION

Complex changes in leukocyte metabolism are associated with the generation of small molecule metabolites, such as ATP, NADPH, nucleotides, and amino acids, which are required rapidly and in high abundance for cell activation, migration and differentiation [1]. Understanding these metabolic networks and adaptations has become more relevant since the discovery of the impact of metabolism on immunity (immunometabolism) and the realisation that cellular metabolism does much more than simply provide energy or biomass. Metabolite and metabolic fluxes can modulate activation or inhibition of cell signalling pathways and post translational modifications (PTMs) to proteins [2].

Dysregulation of immuno-metabolic control has been described in inflammatory diseases including rheumatoid arthritis (RA), where changes in T-cell glycolytic activity drive differentiation, hyper-proliferation and hyper-migration of T-cell subsets [3–5]. RA macrophages have a disease specific metabolic signature which enables CD4+ T cells to differentiate into hyperproliferative pro-inflammatory helper T cells invading tissue and eliciting aggressive tissue inflammation in the synovium through immunogenic cell death [6]. The loss of self-tolerance in people with RA precedes joint inflammation by decades and is a consequence of metabolic dysregulation of both the innate and adaptive immune systems [7].

Neutrophils are the most abundant leukocyte in humans and are specialist cells of the innate immune system that play a major role in host defence against micro-organisms through phagocytosis and generation of ROS [8, 9]. Neutrophils have enormous potential to cause damage to local tissues when dysregulated and are key mediators of inflammation and cartilage damage in RA [10]. Neutrophil survival is enhanced in inflammatory conditions by hypoxia-driven metabolic re-programming [11] and by nutrient availability at the site of inflammation [12]. Neutrophils rely on glycolysis to fuel their energy requirements, where glucose is converted into pyruvate in the cytosol for relatively low-level production of ATP and NADH [13]. The resulting pyruvate is not oxidised in the mitochondria through the TCA cycle, rather it is converted to lactate enabling the generation of NAD+ for re-use in the glycolytic pathway. Furthermore, inflammatory neutrophils contain glycogen, stores of which can be modified when circulating neutrophils are recruited into tissues, by activation of the oxygen-sensing response, and by stimulation with proinflammatory mediators [14, 15]. The first intermediate of glycolysis, glucose 6-phosphate (G6P), also fuels the pentose phosphate pathway (PPP) producing NAPDH which is required to activate the NADPH oxidase (NOX2) leading to ROS production, chromatin decondensation, NOX2-dependent NET formation and NET release [16, 17].

We recently measured the neutrophil metabolome in RA using ^1^H NMR metabolomics and identified lower levels of intracellular glucose and higher levels of lactate and NADP+ in RA neutrophils compared to healthy controls [18]. The reliance on glycolysis for cellular activation is a challenge for neutrophils as sites of inflammation, such as the RA synovial joint, are frequently characterised by limited nutrient availability, including glucose, as well as hypoxia [19]. The aim of this work was to examine the pathways of central carbon metabolism i.e. glycolysis, glycogenolysis, gluconeogenesis and glutaminolysis to determine the importance of these in pathogenic neutrophil release of ROS and NETs in RA.

## 3. METHODS

### 3.1 Isolation and incubation of neutrophils

Ultrapure neutrophils (>99.9% purity) were isolated using the untouched neutrophil isolation kit (Stem Cell). After the blood was incubated 1:5 ratio with Hetasep (Stem Cell) for 30 min at 37°C, the upper layer of nucleated cells was collected and resuspended 1:1 with ice cold isolation buffer (PBS containing 2% BSA and 0.2 mM EDTA). The suspension was then centrifuged at 400g for 5 min and pellet resuspended in 2mL of isolation buffer. Twenty-five microlitres of the antibody cocktail was added to the cell suspension and then incubated for 10 min on ice before adding 50μL of magnetic beads and further incubating for 10 min in an EasySep magnet. Cells were then collected and resuspended in RPMI media (+ L-glutamine, -HEPES, -phenol red) containing either glucose (11mM) or no glucose as indicated at 5×10^6^/mL. In some experiments, neutrophils were pre-treated with 3PO, BPTES, MB05032 or CP-91149 at a range of concentrations (10–50μM), 2-DG (50mM) or AZ-PFKFB3-67 (10nM – 10μM) for 15 min prior to activation with PMA (concentrations below).

### 3.2 Detection of ROS Production By Luminol-Enhanced Chemiluminescence

Following incubation, 2×10^5^ cells were resuspended in 200μL HBSS along with 1μL luminol (final concentration 10μM). The respiratory burst was stimulated with PMA (100ng/mL) and measured in a Fluostar plate reader luminometer at 37 °C.

### 3.3 Measurement of NET Production with Sytox Green

To determine the formation of NETs, 10^5^ neutrophils were seeded in a black 96-well plate containing Sytox green reagent (5μM) and treated firstly with or without cell signalling inhibitors for 15 min in a humidified incubator at 37 °C. Then neutrophils were stimulated with PMA (600nM) and incubated for 4h at 37 °C before fluorescence was measured by Fluostar plate reader at excitation 488nm and emission in the 530nm filter.

### 3.4 Imaging of neutrophils on cover slips

Neutrophils were seeded (at 2×10^5^ cells/500μL) in RPMI media in a 24-well plate containing poly-L-lysine coated coverslips, as previously described [20]. Cells were pre-treated with 3PO (50μM) or left untreated, and allowed to settle and adhere for 30 min prior to stimulation with PMA (600nM). Cells were incubated for a further 4h to allow for NET production. Cells adhered to coverslips were fixed with 4% paraformaldehyde prior to immunofluorescence staining. Briefly, coverslips were removed from the plate and washed with PBS, permeabilised with 0.05% Tween 20 in TBS, fixed with TBS (+2% BSA) and then stained for 30 min on drops of TBS (+2% BSA) on parafilm stretched across a clean 24-well plate. Primary antibodies used were mouse anti-myeloperoxidase (1:1000, Abcam) and rabbit anti-elastase (1:200, Abcam). Coverslips were washed three times with TBS prior to secondary antibody staining (anti-rabbit AlexaFluor488 and anti-mouse AlexaFluor647, 1:2000, Life Technologies) in TBS (+2% BSA) for 30 min. Coverslips were washed prior to staining with DAPI (1μg/mL). Slides were imaged on an Zeiss LSM800 microscope (Zeiss) using a 20X objective.

### 3.5 Measurement of Glycogen content

Neutrophil glycogen content was quantified using the Merck glycogen assay kit. Briefly, 10^6^ neutrophils were lysed in 200μL ice-cold H_2_O and heated for 10 min at 95 °C. Lysates were centrifuged at 18,000g at 4 °C for 10 min to remove cell debris and snap-frozen in liquid nitrogen and stored at -80 °C. On the day of the measurement, 50 μL of defrosted cell lysate was transferred to 96 well black plate and 1μL of hydrolysis enzyme mix was added and the plate incubated for 30 min at room temperature. To remove the effect of background glucose, a sample blank was set up for each reaction by omitting the hydrolysis enzyme mix. The sample blank was then subtracted from the sample readings. After incubation, 48μL of development buffer was added to each well, with 1μL of development enzyme mix and 1μL of fluorescent peroxidase substrate. The plate was incubated for 30 min at room temperature and protected from light. Fluorometric measurements were carried out with Fluostar plate reader with an excitation of 535nm and emission of 587nm. The final concentration of glycogen in the samples was calculated from the standard curve as per the manufacturer’s instructions.

### 3.6 Measurement of OCR with Seahorse

Oxygen consumption rate (OCR) was measured using the Seahorse XFe96 Analyser (Agilent Technologies). Neutrophils (2.5×10^5^ per well) were seeded onto XFe96 cell culture microplates (Agilent Technologies) pre-coated with Cell-Tak. RPMI medium was supplemented with 2mM glutamine, 10mM glucose, 1mM pyruvate (all from Agilent Technologies). Plates were incubated for 30 min at 37°C and loaded into the Seahorse XFe96 Analyser. OCR was quantified at the beginning of the assay and after the sequential injection of oligomycin (2μM), Carbonyl cyanide-4-(trifluoromethoxy)phenylhydrazone (FCCP, 0.5μM) and rotenone plus antimycin A (0.5μM) (MitoStress Test kit, Agilent Technologies). PMA (600nM) was injected after 15 min to stimulate the oxidative burst which was quantified by calculating the area under the curve. Assays were performed with 5 technical replicates from each neutrophil donor, with OCR plotted with standard error and mean for each time point.

### 3.7 Measurement of Apoptosis By Flow Cytometry

Following incubation with or without inhibitors for up to 24h, 5×10^4^ neutrophils were removed from culture and diluted with 200μL HBSS containing 0.5μL Annexin V-488, and incubated at room temperature in the dark for 15 min. The total volume was then made up to 250μL with HBSS, and 0.25μL propidium-iodide added (1μg/mL, final conc) before reading on a EasyCyte Guava flow cytometer. Five thousand events were analysed per sample.

### 3.8 Measurement of GLUT3 by flow cytometry

Neutrophils (5×10^4^) were labelled with saturating concentrations of FITC-labelled anti-GLUT3 antibody or isotype control (BD Biosciences). Unlabelled cells were also prepared as a control. Cells were incubated in PBS (0.2% BSA) with or without antibodies for 30 min at 4°C. Cells were washed in PBS (BSA) and then fixed using 4% (final conc) paraformaldehyde in PBS. Cells were washed again to remove excess paraformaldehyde, and then resuspended in 200μL PBS. Cell surface antibodies were detected using a Cytoflex Beckman Coulter 2.0 flow cytometer, with 10,000 cells counted per sample.

### 3.9 Gene expression analysis

Neutrophil RNA was isolated by Trizol and cleaned with the Qiagen RNeasy kit including on-column DNase digestion. RNAseq was performed using standard protocols by BGI Tech Solutions (Hong Kong Ltd). RNAseq datasets were analysed by DESeq2 [21] to detect genes significantly different between RA (n=65) and HC (n=11) (adj. p-value < 0.05). Gene ontology analysis was carried out on genes with a 1.5 fold change in gene expression between RA and HC neutrophils using clusterProfiler [22, 23] and enrichPlot [24] in R v4.0.

### 3.10 NMR Metabolomics

Neutrophils were prepared for metabolite extraction as previously described [18]. Briefly, cells were centrifuged at 1000g at 25°C for 2 min, the pellet washed with ice cold PBS then centrifuged at 1000g at 25°C for 2 min. The supernatant was discarded, while pellets were heated at 100°C for 1 min, and then snap-frozen in liquid nitrogen and stored at -80°C. Metabolites were extracted by addition of 50:50 v/v ice cold HPLC grade acetonitrile:water at 500μL per cell pellet, followed by a 10 min incubation on ice. Samples were sonicated while incubated in an ice water bath for three 30 s bursts at 23 kHz and 10μm amplitude using an exponential microtip probe, with 30s rest between sonications. Sonicated samples were centrifuged at 12,000g for 5 min at 4°C and the supernatant transferred to cryovials, flash frozen in liquid N_2_ and lyophilised prior to storage at -80°C. Each lyophilised sample was resuspended in 200μL of 100μM deuterated sodium phosphate buffer pH 7.4, with 100μM trimethylsilyl propionate (TSP) and 0.05% NaN_3_. Each sample was vortexed for 20s and centrifuged at 21500g for 5 min at 5°C. Then 180μL of each cell extract sample was transferred to 3 mm (outer diameter) NMR tubes for acquisition. The samples were analysed using a 700 MHz NMR Avance IIIHD Bruker NMR spectrometer equipped with a TCI cryoprobe. Samples were referenced to trimethylsilylpropanoic acid (TSP) at 0 ppm. Spectra were acquired at 25°C using the 1D ^1^H Carr–Purcell– Meiboom–Gill (CPMG) edited pulse sequence (vendor supplied cpmgpr1d) with 512 scans and a 4s interscan delay and 1.8s acquisition time. Spectra were proceed using vendor supplied auto routine (apk0.noe) prior to manual acceptance via quality control criteria. Spectra were assessed to conform to minimum quality criteria as outlined by the Metabolomics Standards Initiative [25] to ensure linewidths at half height within one standard deviation with consistent signal-to-noise, baseline correction and water suppression. All spectra passing quality criteria were then divided into peak boundaries “bins” that were defined globally by the peak limits using Chenomx NMR Suite 8.2 (Chenomx Inc., Edmonton, Alberta, Canada)[26]. All peaks, both annotated in Chenomx (via manual analyses in TopSpin and Chenomx software) and unknowns, were included in the bin table. A correlation-reliability scoring (CRS) method comparing metabolite annotated peaks via a Pearson correlation [18, 27] was applied to the data which aimed to address the problem of selecting appropriate representative bins from feature extraction in multivariate analysis [27].

### 3.11 Preparation of immune complexes

Insoluble immune complexes (IIC) were prepared by incubating human serum albumin with anti-human serum albumin antibody in PBS for 1h as previously described [28]. Immune complexes were washed 3 times by centrifugation at 1,000g and re-suspended in PBS before use to remove soluble immune complexes. In some experiments neutrophils were pre-treated with 3PO (0-50μM) for 15 min prior to stimulation with 10% (v/v) IIC to stimulate NET production.

### 3.12 Western Blotting

Neutrophils were boiled in Laemmli buffer containing 50mM DTT at 95°C for 5 min. Proteins were separated by SDS-PAGE using a NuPAGE bis-Tris 4–12% gradient gel and transferred into a PDVF membrane using the Trans-Blot Turbo apparatus (Bio-Rad). Membranes were blocked in 5% BSA in Tris-buffered saline-Tween 20 (TBS-T), prior to overnight incubation at 4°C with primary antibodies: PFKFB3 (1:1000, Abcam) and Actin (1:10,000, Abcam). Blots were then washed in TBS-T prior to incubation with anti-rabbit IgG HRP secondary antibody (1:10000, Cell Signaling) and anti-mouse IgG HRP secondary antibody (1:10000, Sigma). Bands were detected using enhanced chemiluminescent substrate (ECL; Thermo Fisher, Loughborough, UK) and imaged on a ChemiDoc image analyser (Bio-Rad Laboratories).

### 3.13 Bacterial killing assay

Bacterial killing assay was performed as previously described [29]. Briefly, freshly-grown *S. aureus* were harvested and washed, and suspended at 5×10^8^/ml in HBSS and opsonised with 10% human AB serum (Sigma) for 30 min at 37°C. Freshly-isolated neutrophils (10^6^/mL) were primed with TNFα (10ng/mL) and incubated for 1h at 37°C with gentle agitation with serum-opsonised bacteria at a ratio of 1:10. Neutrophils were then lysed to release live bacteria by serial dilution in distilled water and vigorous vortexing, before being plated onto LB agar plates and incubated overnight. Colonies were counted and results calculated as percentage of bacteria killed compared to bacteria only (no neutrophils) samples.

### 3.14 Data analysis

Statistical analyses were performed using R v4.0.2. For experimental data, the Shapiro-Wilk test was used to determine normality. Then univariate analysis was carried out by the Student’s t-test, Wilcox test or ANOVA as appropriate. Multivariate ^1^H NMR metabolomics data was assessed for normality via qqplots, and found to be non-normally distributed therefore a Wilcox test was performed with application of a False-Discovery Rate (FDR) and adjusted p-value of 0.05. Fold change comparisons were performed comparing natural log of median metabolite abundances on probabilistic quotient normalised data. Metabolic pathway enrichment analysis was performed using fold change analysis to select 15 metabolites with mean fold change greater than 20% and metabolite set enrichment analysis against the small molecule pathway database (SMPD) of human metabolic pathways using Metaboanalyst v6.0 [30].

## 4. RESULTS

### 4.1 Increased expression of genes regulating central carbon metabolism in RA neutrophils

In our previous study we identified that the metabolome in RA neutrophils is different from HC, mainly driven by metabolites of central carbon metabolism (glucose, ATP, NADP, lactate) [18]. In this study we wanted to identify the underlying mechanisms responsible for these differences, and therefore we first analysed the expression of genes regulating glycolysis in RA neutrophils. Using RNAseq, we identified gene ontology pathways significantly enriched in RA neutrophils compared to HC. This identified increased metabolic pathways in RA neutrophils, including: ‘cellular response to glucose starvation’; ‘fructose metabolic process’ (p<0.05) and ‘glycolytic process through fructose-6-phosphate’ (p>0.05). We also identified up-regulation of neutrophil effector functions including: ‘positive regulation of cytokine production’; ’regulation of leukocyte cell-cell adhesion’; ‘response to molecule of bacterial origin’; ‘response to interferon gamma’ and ‘hydrogen peroxide metabolic process’ (p<0.05, Figure 1A). We then analysed the expression of specific genes involved in glycolysis in RA and HC neutrophils. The gene for 6-Phosphofructo-2-Kinase/Fructose-2,6-Bisphosphatase 3 (PFKFB3) was significantly increased in RA (adj. P-value = 0.01, Figure 1B). The other related genes, 6-Phosphofructo-2-Kinase/Fructose-2,6-Bisphosphatase 2 (PFKFB2) and 6-Phosphofructo-2-Kinase/Fructose-2,6-Bisphosphatase 4 (PFKFB4) that together form the isoenzyme of Phosphofructokinase-2 (PFK2), were not significantly different (Figure 1B). Neutrophil transcriptomics analysis also highlighted an increase in gene expression for the glucose transporter gene SLC2A3 which codes for the glucose transporter-3 (GLUT3) protein (Figure 1B, adj. P-value = 0.04) in RA neutrophils. We confirmed increased plasma membrane expression of GLUT3 on RA neutrophils by flow cytometry (Figure 1C, p<0.05), and increased total protein expression of PFKFB3 in RA was confirmed by Western Blotting (Figure 1D).

**Figure 1.**
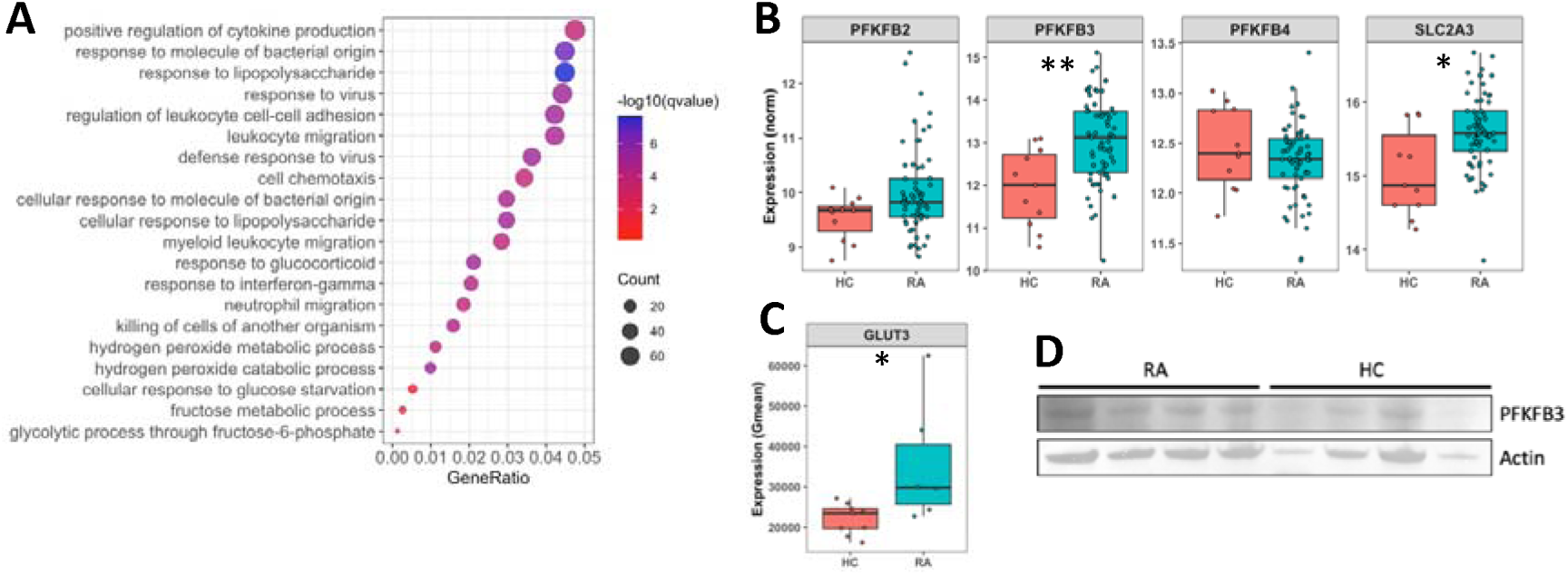
Dysregulation of metabolic gene expression in RA neutrophils. (A) Gene ontology analysis of genes expressed >1.5 fold higher in RA compared to HC. (B) Boxplots showing expression levels of PFKFB2, PFKFB3, PFKFB4, SLC2A3 in RA and HC neutrophils (*FDR<0.05, **FDR<0.01). (C) GLUT3 protein on the surface of HC and RA neutrophils (*p<0.05). (D) Western blot of PFKFB3 and Actin in freshly isolated RA and HC neutrophils (each n=4).

### 4.2 Absence of extracellular glucose does not change ROS or NETs in RA neutrophils, but inhibition of glycolysis blocks ROS and NET production in both HC and RA neutrophils

In order to investigate the requirement for glycolysis to fuel ROS and NET production we incubated HC and RA neutrophils in media containing 11mM glucose (glucose-containing) or media without glucose (no glucose) media in the presence of PMA and measured ROS continuously by luminol-enhanced chemiluminescence. The absence of glucose in the media did not affect neutrophil ROS production (Figure 2A) but did decrease NET production by HC neutrophils (Figure 2B, p<0.01). In contrast, RA neutrophil NETs were not affected by the absence of extracellular glucose, indicating an adaptation to preserve ROS production in low-glucose environments. Competitive inhibition of the first step of glycolysis (the production of glucose-6-phosphate by hexokinase) using 2-deoxy-D-glucose (2-DG) significantly decreased ROS production by both HC and in RA neutrophils (Figure 2C, p-value<0.01). 2-DG also completely inhibited NET production in glucose-containing or no glucose media in both HC and RA neutrophils (Figure 2D, p-value<0.01).

**Figure 2.**
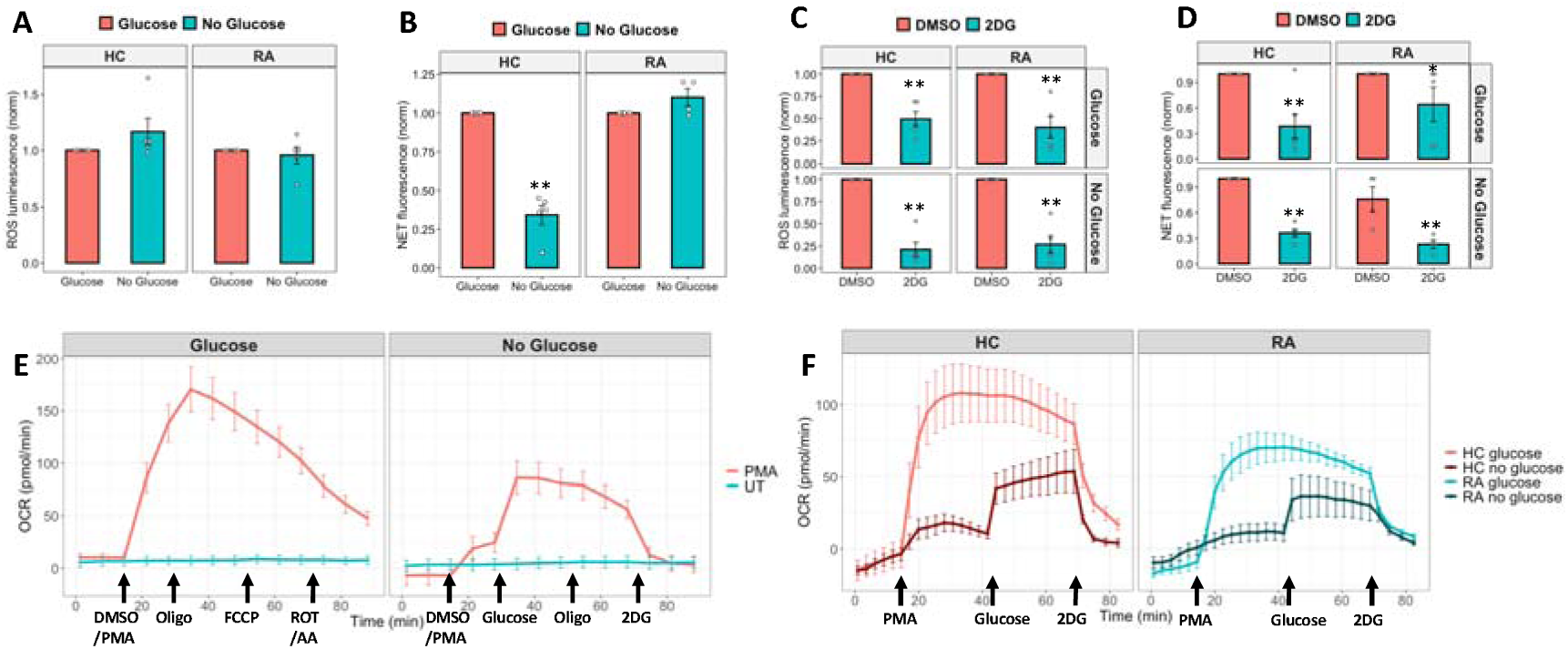
Effect of glucose availability on ROS and NET production. ROS (A,C) and NET (B,D) release in response to PMA in glucose-containing or no glucose media in the absence (A,B) or presence (C,D) of 2-DG (* p<0.05, **p<0.01). (E) Seahorse XF OCR trace of HC neutrophils incubated in glucose-containing or no glucose media. Neutrophils were activated by injection of PMA (600nM) into the media after 15 min as indicated. Neutrophils in glucose-containing media were challenged with mitochondrial inhibitors: oligomycin (2μM), FCCP (0.5μM), and antimycin A plus rotenone (Rot/AA, 0.5μM) at timepoints indicated by arrows, whereas neutrophils in no glucose media were injected with glucose (11mM), oligomycin (2μM) and 2-DG (50μM) at timepoints indicated by arrows (n = 2 biological replicates, n = 5 technical replicates). (F) Seahorse XF OCR trace of HC and RA neutrophils incubated in glucose-containing or no glucose media. Neutrophils were activated with PMA after 15 min as indicated by an arrow and 2-DG was injected where indicated. Neutrophils in no glucose media were given glucose as indicated.

We next used the Seahorse XF assay to determine the changes in oxygen consumption rate (OCR) induced by either inhibition of glycolysis (by 2-DG) or the absence of glucose in the media, with and without PMA stimulation. Non-activated (untreated) neutrophils did not show any change in OCR in response to oligomycin (ATP synthase complex V inhibitor), FCCP (uncoupler of mitochondrial oxidative phosphorylation) or Rotenone and Antimycin (Rot/AA, complex I and III inhibitors) showing that resting human neutrophils do not use mitochondrial respiration to produce ATP (Figure 2E). PMA activation on the other hand stimulated an increase in OCR by neutrophils. However after a second injection of oligomycin, no decrease in OCR was observed after the initial OCR spike. PMA-activated neutrophils were also unaffected by FCCP or Rot/AA, again showing that activated neutrophils do not utilise mitochondrial respiration, and indicating that the increase in OCR detected was caused by formation of ROS via NOX2. When neutrophils were incubated in media without glucose, PMA-activated neutrophils still consumed oxygen, however at a lower rate than in glucose-containing media. Injection of glucose into the media after 30 min augmented OCR. Injection of 2-DG completely decreased OCR by activated neutrophils highlighting the requirement for glycolysis in NOX2-dependent ROS production (Figure 2F).

### 4.3 Glycogen is stored by neutrophils in low glucose environments

Glucose may be stored by neutrophils in the form of glycogen, a multi-branched polysaccharide of glucose molecules, in order to fuel glycolysis when extracellular sources cannot meet the metabolic demands of the cell. We therefore measured the glycogen content in HC and RA neutrophils after incubation for 1h in glucose-containing or no glucose media. Glycogen stores increased when glucose was absent from the media in both HC and RA neutrophils, but this increase was only statistically-significant in HC neutrophils (Figure 3A, p-value<0.05). The increase in intracellular glycogen levels in media without glucose was inhibited when neutrophils were treated with 2-DG (Figure 3A, p-value<0.05). To determine whether glycogen is utilised by neutrophils to provide the G6P needed to fuel ROS and NET production via the Pentose Phosphate Pathway (PPP), neutrophil activation was measured in the absence and presence of CP-91149 (0–50μM), a selective inhibitor of glycogen phosphorylase. No significant differences in ROS or NET production were observed in the presence of CP-91149 in HC and RA neutrophils incubated in glucose-containing or no glucose media. (Figure 3B,C).

**Figure 3.**
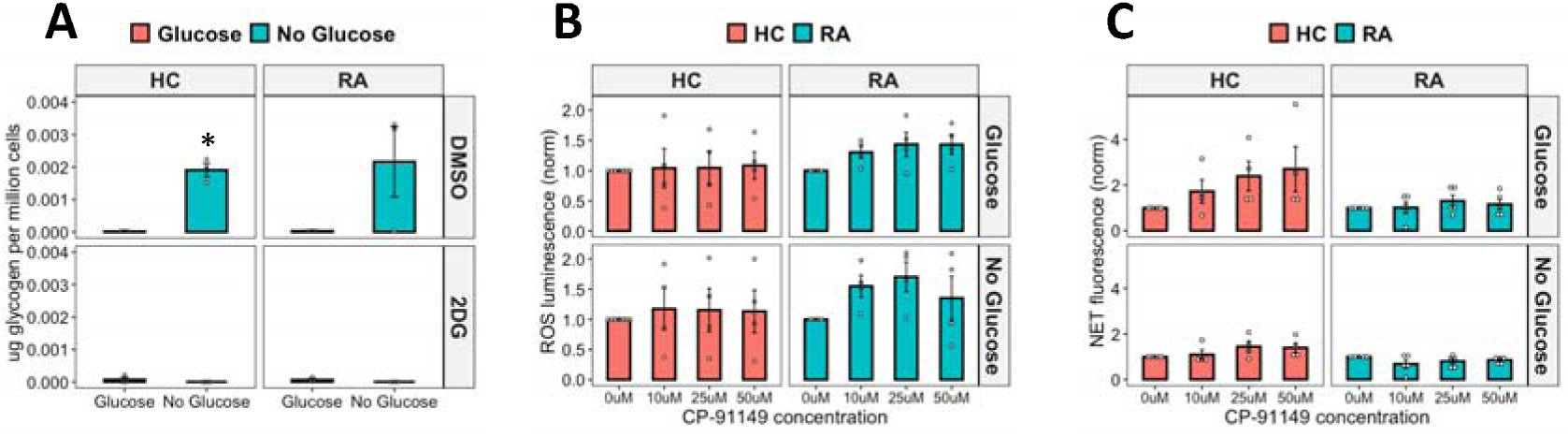
Effect of extracellular glucose on glycogen stores in HC and RA neutrophils. (A) Measurement of intracellular glycogen content of neutrophils incubated in glucose-containing or no glucose media and treated with or without 2-DG for 1h. (* p<0.05). (B) Effect of inhibiting glycogenolysis on neutrophil (B) ROS and (C) NET production by neutrophils incubated in glucose-containing or no glucose media and treated with increasing concentrations of inhibitor of glycogenolysis CP-91149 (0 – 50 ∝M).

### 4.4 The chemical inhibitor 3PO blocks ROS and NET production by RA and HC neutrophils

Fructose-2,6-phosphate (PFK-2) is a key enzyme in the early stages of glycolysis, and our gene expression analysis identified that the PFK-2 subunit PFKFB3 was significantly increased in RA neutrophils (Figure 1B). We therefore inhibited PFK-2 activity using 3PO, a chemical inhibitor of the PFKFB3 subunit. ROS production was inhibited in a dose-dependent manner (0-50μM) in both HC and RA neutrophils (Figure 4A, p<0.01). In HC and RA neutrophils, increasing concentrations of 3PO significantly decreased ROS production in both types of media. RA neutrophil NET production was also significantly inhibited at all concentrations from 10μM to 50μM (p<0.001). However, in HC neutrophils, 3PO did not inhibit NETs at a concentration of 10μM, and a significant decrease was only observed at the higher concentrations of 25μM and 50μM (Figure 4B,C p<0.01) indicating RA neutrophils may be more sensitive to 3PO treatment. This effect was confirmed by immunofluorescence staining of NETs on coverslips after 4h incubation with PMA in the absence or presence of 3PO (50μM) (Figure 4C). We also determined that 3PO significantly inhibited NET production at all concentrations in RA and HC neutrophils in response to immune complexes (IIC) which mimic auto-antibody immune complexes such as rheumatoid factor, commonly found in the serum and synovial fluid of people with RA (Figure 4D, p<0.01). Finally when neutrophils were pre-treated with 3PO (0-50μM) we observed a dose-dependent decrease, but not complete inhibition, of bacterial killing capacity (Figure 4E, p<0.05).

**Figure 4.**
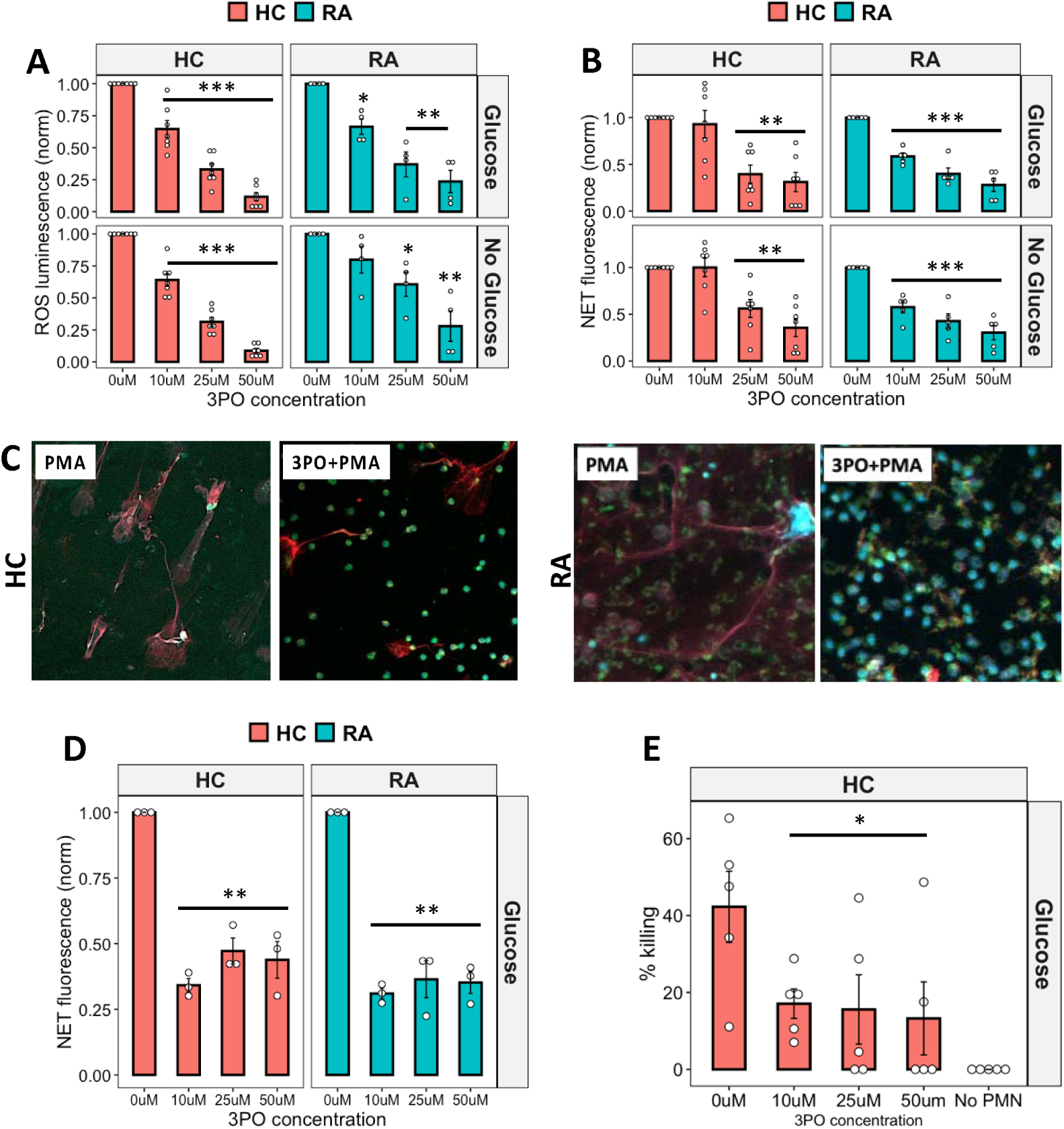
Anti-inflammatory effect of 3PO on neutrophils. (A) ROS (B) and NET production in response to PMA by neutrophils incubated with 3PO (0 – 50 ∝M) in glucose-containing or no glucose media (* p<0.05, **p<0.01, ***p<0.001). (C) Representative images of HC and RA NETs produced in response to PMA in the absence or presence of 3PO (50∝M). Blue = DNA, DAPI, Red = MPO, Green = elastase. (D) NET production in response to insoluble immune complexes incubated with 3PO (0 – 50 ∝M, **p<0.01). (E) Killing of S. aureus by 3PO-treated neutrophils (0 – 50 ∝M, *p<0.05) compared to no neutrophil (No PMN) control.

Next we determined the effect of 3PO on neutrophil apoptosis by incubating RA and HC neutrophils for 24h in the absence and presence of 3PO. Interestingly, in both glucose-containing and no glucose media we observed no difference in the overall percentage of live (annexin V negative, PI negative) neutrophils. However the populations of early apoptotic (annexin V positive, PI negative) and late apoptotic (annexin V positive, PI positive) cells were significantly decreased and increased respectively in both RA and HC by 3PO (Figure 5A, p<0.05).

**Figure 5.**
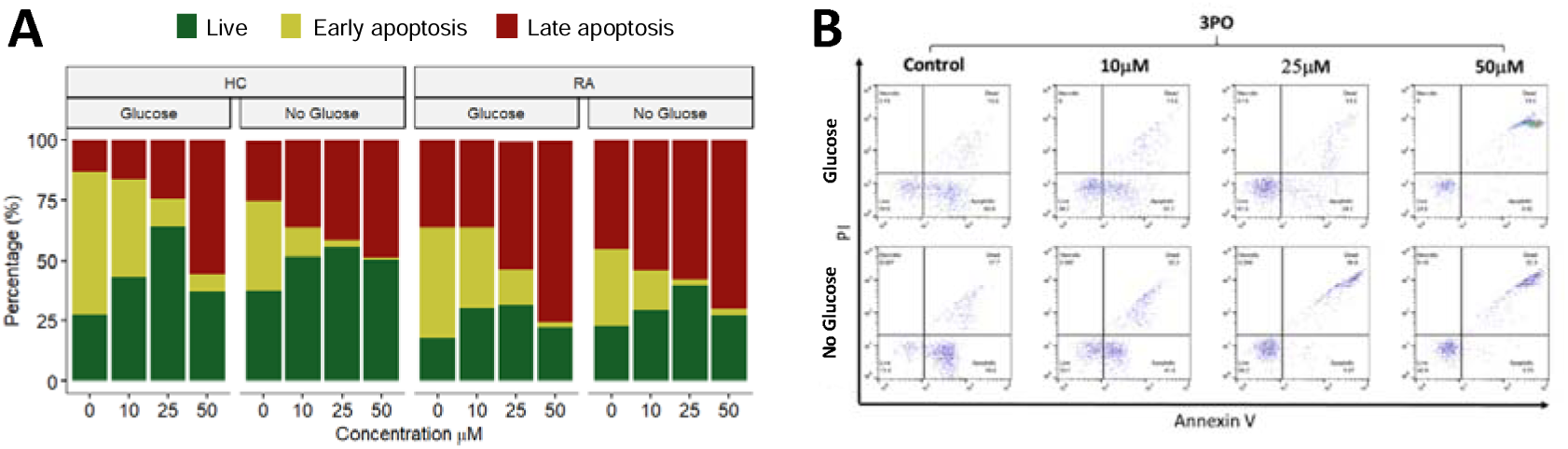
Effect 3PO on neutrophil apoptosis. (A) Percentage of late apoptosis (red), early apoptosis (yellow) or live (green) HC and RA neutrophils (n=3 each) incubated in glucose or no glucose media and treated for 24 h with varying concentrations of 3PO (0 – 50 ∝M). (B) Representative flow cytometry traces for HC neutrophils showing annexin V and PI positive/negative gating.

### 4.5 3PO may exert an off-target anti-inflammatory effect in human neutrophils

Recent reports on the use of 3PO to inhibit PFKFB3 have shed doubt on the specificity of this compound to bind to its reported target [31, 32]. Indeed recent publications have suggested that whilst 3PO does inhibit glycolysis, this is not in response to PFKFB3 binding and inhibition [31, 32]. Off-target inhibition of NF-κB by 3PO has also been reported [32]. We therefore decided to repeat the ROS and NET assays using an alternative inhibitor of PFKFB3, AZ-PFKFB3-67 (AZ67). This compound is reported to be a highly specific inhibitor of PFKFB3 with an IC_50_ of 11nM. We found that pre-treatment of HC neutrophils with increasing concentrations of AZ67 (10nM – 10μM) had no effect on neutrophil ROS or NET production (Figure 6A,B), in contrast to our results with 3PO.

**Figure 6.**
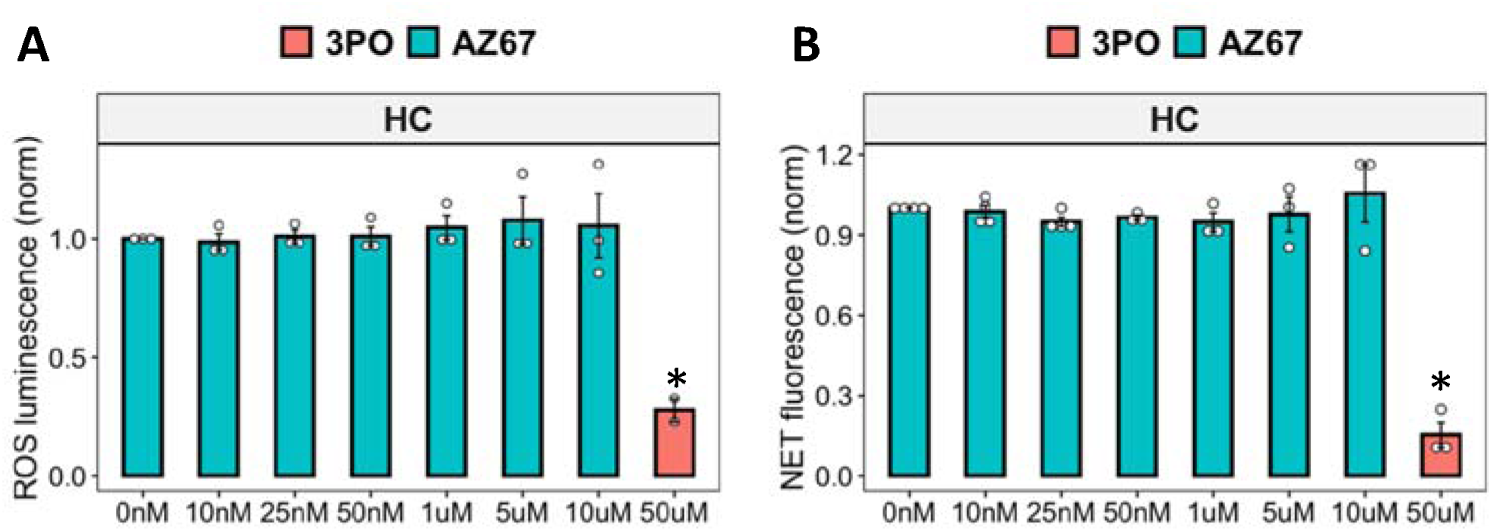
Inhibition of PFKFB3 in neutrophils using AZ67. (A) ROS (B) and NET production in response to PMA by neutrophils incubated with AZ-PFKFB3-67 (0 – 10 ∝M) in glucose media, with 3PO (50 ∝M) as positive control (* p<0.05).

In order to determine the molecular mechanism of ROS and NET inhibition by 3PO we performed ^1^H NMR metabolomics analysis on neutrophils incubated for 4h in the absence or presence of 3PO (50μM) in glucose-containing media. We found that 3PO significantly decreased the levels of glutathione in RA and HC neutrophils (Figure 7A, adj.p<0.05). Mean L-glutamine and phenylalanine levels increased by 1.9-fold and 1.35-fold respectively, although this was not statistically significant (Figure 7A). Mean levels of lactate increased by 1.5-fold after 3PO treatment (Figure 7A). Pathway enrichment analysis of metabolites that increased or decreased by at least 20% in response to 3PO was performed using MetaboAnalyst and is summarised in Figure 7B. Pathways that were significantly over represented in metabolite variance associated with 3PO included those relating to membrane lipid synthesis, glutathione metabolism and amino acid recycling.

**Figure 7.**
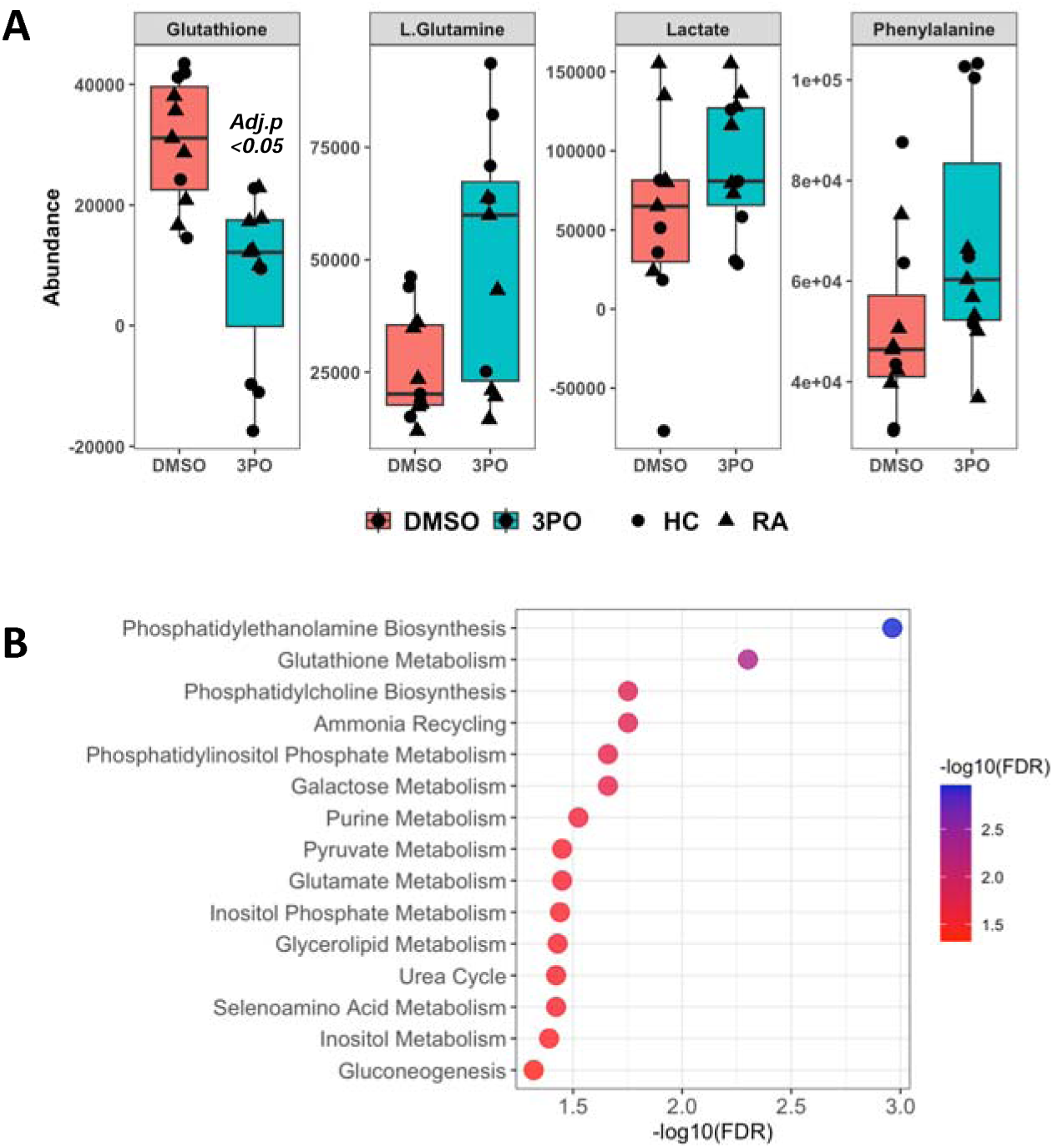
^1^HNMR metabolomics analys is of RA and HC neutrophils with PFKFB3 inhibition by 3PO. (A) Boxplots of metabolites measured by ^1^H-NMR from neutrophils incubated in glucose media for 4h with and without treatment with 50∝M 3PO. Glutathione was significantly lower in 3PO-treated neutrophils (adj,p-value<0.05). (B) Pathway enrichment analysis results showing pathways with at least 3 metabolites and FDR-adjusted p-value < 0.05.

### 4.6 Inhibitors of gluconeogenesis and glutaminolysis do not affect ROS and NET production in RA and HC neutrophils

Finally we investigated the effect of inhibiting gluconeogenesis and glutaminolysis on ROS and NET production by RA and HC neutrophils. MB05032 is a potent FBPase (FBP1) inhibitor which inhibits gluconeogenesis and BPTES inhibits glutaminase-1. MB05032 inhibited HC NETs at the highest concentration tested (50μM, p<0.05) but otherwise neither inhibitor significantly inhibited ROS or NET production in RA or HC neutrophils in glucose-containing or no glucose media (Figure 8).

**Figure 8.**
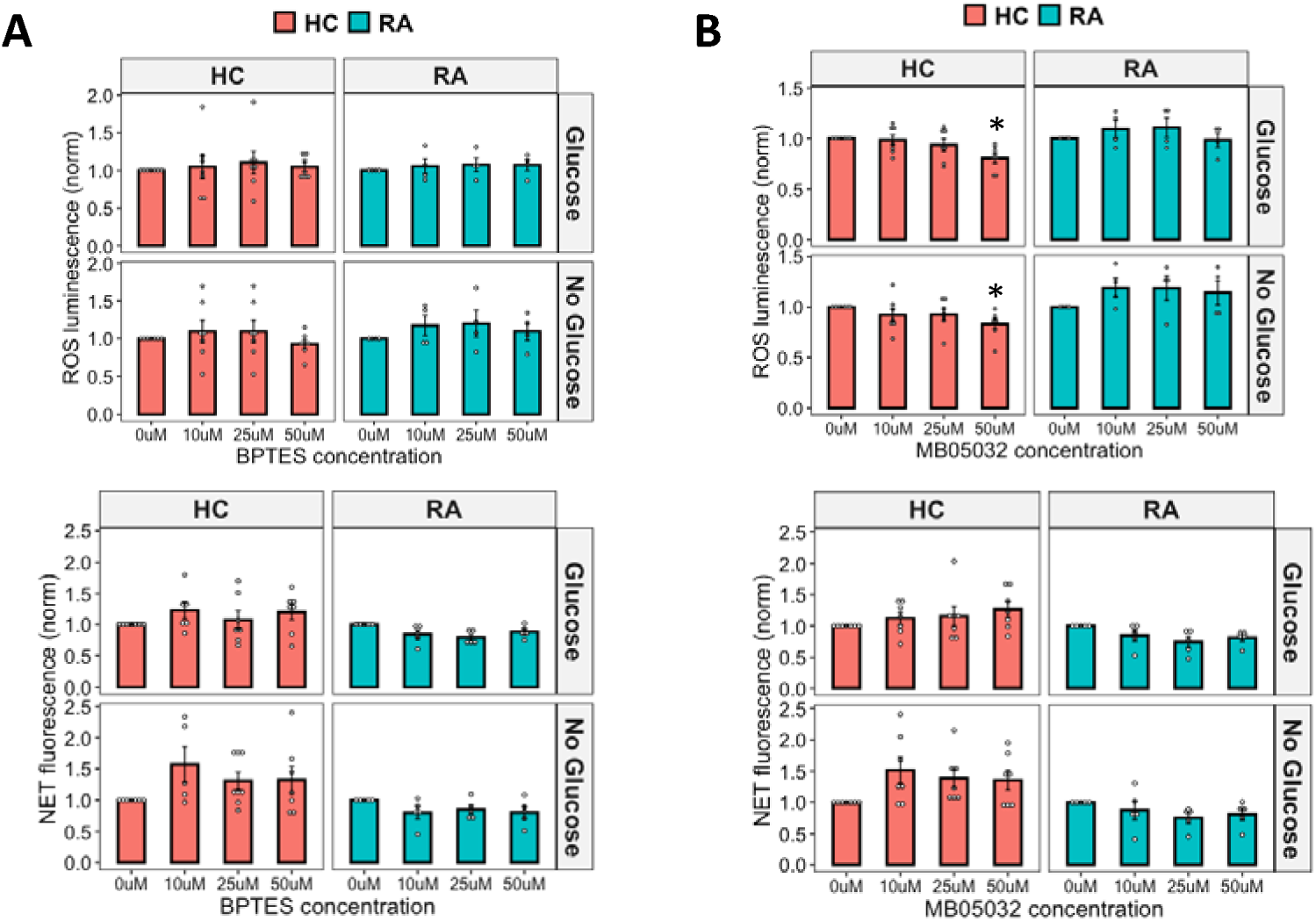
Effect of glutaminolysis and gluconeogenesis on neutrophil activation. (A) ROS (B) and NET production in response to PMA by RA and HC neutrophils in glucose-containing or no glucose media when treated with an inhibitor of gluconeogenesis MB05032 or an inhibitor of glutaminolysis BPTES (0 – 50 mM) (*p<0.05).

## 5. DISCUSSION

In this study we measured HC and RA neutrophil responses when challenged with inhibitors of central carbon metabolism or low extracellular glucose concentrations. Recent studies have highlighted the importance of pathways connected to glycolysis in neutrophils, including glycogenolysis, gluconeogenesis, glutaminolysis and PPP [17, 18] and this work provides new information about the critical role of glucose metabolism in regulating ROS production, NET release and apoptosis in human neutrophils.

Neutrophils are the first responders to inflamed sites. These environments generally have lower glucose concentrations than peripheral blood, a potentially unfavourable environment for neutrophils given their reliance on glycolysis for energy production and activation [16]. However, neutrophils are often found at high concentrations in low glucose, inflamed sites, for example in synovial fluid from people with RA [19]. Our first experiments confirmed the reports of others [33–35] that mitochondrial respiration is not required for energy production or an oxidative burst in human HC or RA neutrophils. This notion was recently challenged by a study which suggested that the glucose-restricted tumour microenvironment induces the formation of immature murine, c-Kit+ neutrophils subsets which maintain local immune suppression by using mitochondrial fatty acid oxidation to support NADPH oxidase-dependent ROS production [34]. We believe this highlights another important difference between human and murine neutrophils [36] underlying the importance of our use of human neutrophils in this study. Using the Seahorse XFe96 XF analyser we showed that when HC and RA neutrophils were challenged with inhibitors or stimulators of mitochondrial respiration there was no change in OCR. This demonstrates that mitochondria are dispensable for ATP production in human neutrophils.

We explored neutrophil ROS production using a combination of luminol-enhanced chemiluminescence and the Seahorse XFe96 XF analyser. The luminol-enhanced chemiluminescence assay is relatively simple to use and is a high sensitivity and cost effective method to detect ROS production and measures both extracellular and intracellular ROS [37]. ROS production measured by Seahorse XFe96 XF provides a direct, non-invasive, real-time detection and quantification of neutrophil activation by measuring oxygen consumption rate (OCR). We confirmed that neutrophils do require glycolysis for ROS and NET production, given the complete inhibition of both in HC and RA neutrophils by 2-DG as previously reported. However, the absence of glucose in the extracellular media did not completely inhibit ROS production. Indeed, we detected decreased OCR of activated neutrophils when measured by Seahorse compared to neutrophils in glucose-containing media. However, no decrease was observed when neutrophil ROS was measured by luminol-enhanced chemiluminescence in depleted media. These diverging results could be attributed to the mode of ROS detection by the two different assays. The Seahorse OCR assay measures depletion of O_2_ in the culture media, in this case via the activity of the NOX2 NADPH oxidase. This is the first step in ROS production. During the respiratory burst, NOX2 reduces superoxide to ^2^O_2-_, which is then converted to hydrogen peroxide (H2O_2_) by superoxide dismutase. Hydrogen peroxide may then react with MPO to form hypochlorous acid (HOCl) and other secondary oxidants [8]. Luminol detects both extracellular and intracellular ROS production by NOX2 and MPO. NET formation is dependent on ROS production and inhibition of intragranular MPO activity has been shown to correlate with inhibition of NET formation [37–39]. When measuring NET formation at a single timepoint of 4h, no differences were observed between glucose-containing and no glucose media. However complete inhibition of glycolysis by 2-DG prevented NET formation. This could highlight that intracellular ROS formation, irrespective of the extracellular glucose environment, is sufficient for processing by MPO in order to trigger NET formation.

Glycogen stores may be broken down to increase intracellular glucose levels, and as previously shown glycogen is essential for inflammatory neutrophil function [12]. Neutrophils accumulate glycogen as they transition from circulating to elicited peritoneal cells in guinea pigs [15]. A more recent study observed an increase in glycogen granules during inflammation in neutrophils from people with sepsis [40], and the authors suggested the morphological evaluation of the cells could be a useful biomarker for rapid and economical diagnosis of sepsis. The increase in glycogen stores has also been studied in Covid-19 patients where neutrophils have an increased amount of stored glycogen compared to healthy controls [41]. In our experiments we observed a significant increase in glycogen storage when both HC and RA neutrophils were incubated in no glucose media for 1h, which highlights a metabolic adaptation to the extracellular environment. The no glucose media used in our experiments mimics the low glucose environment found within inflamed RA synovial joints [19], with the increase in glycogen stores reflecting a metabolic switch to an inflammatory phenotype. Inhibition of glycolysis by 2-DG completely stopped glycogen formation. This is because the first step in glycogen formation requires the transformation of glucose to glucose-6-phosphate by hexokinase, a reaction that is inhibited when 2-DG forms 2-deoxy-D-glucose-phosphate. When ROS production was measured in the presence of the inhibitor of glycogenolysis, CP-91449, no significant change in ROS or NET production by HC or RA neutrophils was measured.

Our transcriptomics data provided an extra layer of insight into the dysregulation of metabolic pathways in RA neutrophils. Functional enrichment analysis identified glycolytic processes as being upregulated in RA neutrophils, with the most significantly altered gene in RA being PFKFB3. Four different PFK-2/FBPase-2 isozymes have been identified and the overexpression of two isozymes (PFKFB3 and PFKFB4) has been demonstrated in various solid tumours and haematological cancer cells [42]. Peripheral and tissue resident T-cells from people with RA have a unique metabolic signature with an impairment of glycolysis due to a deficiency of PFKFB3, resulting in delayed glycolysis and increased PPP via the up-regulation of glucose-6-phosphate dehydrogenase (G6PD) [3]. The ratio of these two enzymes in T cells correlates to disease activity in people with RA which suggests a dysregulation of pro-inflammatory properties of T cells directly affected by PFKFB3 and G6PD [4, 43]. T cells are therefore diverting glucose from energy generation towards synthesis of biomass precursors with functional consequences that include hyper-proliferation, G2/M bypass and deviated functional commitment [44].

Little is known about the role of PFKFB3 in regulating neutrophil metabolism and function. Enhancement of PFKFB3 transcription and translation facilitates the production of neutrophil inflammatory factors during the acute phase of sepsis [45]. Pharmacological inhibition or depletion of PFK-2 by small interfering RNA (siRNA) leads to a significant decrease in NOX2 activity response to various neutrophil stimuli [46]. Among the four isoenzymes of PFK-2, PFKFB3 and PFKFB4 are the two main isoenzymes overexpressed in various human cancers and in neutrophils. PFKFB3 is a critical glycolysis regulatory enzyme that promotes fructose 2,6-bisphosphate (F2,6BP) production which in turn activates PFK-1, increasing glycolytic output. It has the highest kinase:phosphatase activity ratio of all the PFKFB isoforms [47], proving a possible explanation as to why inhibition of this isoform has such a marked effect on cellular function. In vitro experiments showed that glycolytic metabolism with PFKFB3 involvement supports inflammatory cytokine expression [45]. Furthermore, a significant increase has been observed in the expression of PFKFB3 in LPS-challenged and sepsis neutrophils compared to controls [48]. PFKFB4 is associated with immune cell infiltration and immunological checkpoints in non-small cell lung cancer [49]. In neutrophils, selectively targeting phosphofructokinase-1 liver type (PFKL) suppresses the NOX2-dependent oxidative burst [50]. It was also observed that PMA, LPS, or cholesterol crystals decreased PFKL activity to promote NOX2-dependet NETosis, except when the cells were treated with PFKL agonists NA-11 or LDC7559. In contrast, NETosis induced in a NOX2-independent manner by the ionophore nigericin did not change PFKL activity and was not blocked by NA-11 [50]. PFKL knockdown in macrophages also decreased glycolytic flux as evidenced by diminished lactate and resulted in diversion of glucose into the PPP [51]. However, little else is known about the role of phosphofructokinase enzymes in neutrophils. For this reason we tested whether neutrophils were still able to activate the NOX2 pathway to produce ROS and NETs when challenged with the PFKFB3 inhibitor 3PO. We observed a concentration-dependent inhibition of ROS production by 3PO in HC and RA neutrophils in both glucose-containing and no glucose media. NET production was also inhibited by 3PO, although RA neutrophils were more sensitive to 3PO inhibition at lower concentrations than HC neutrophils. Importantly, 3PO inhibited NET production in response to immune-complexes (IIC) which mimic the auto-immune antibodies found in the majority of people with RA. Measurement of neutrophil apoptosis after 24h showed that increasing concentrations of 3PO increased the number of late apoptotic cells and decreased the early apoptotic cell count. The increase in late apoptotic cells at the highest concentrations of 3PO may be undesirable as these cells have leaky membranes and are likely to be pro-inflammatory especially at inflammatory sites. We also observed that bacterial killing was suppressed by 3PO. However, bacterial killing was not completely abrogated even at relatively high 3PO concentrations, indicating that the neutrophils retained some phagocytosis and killing functions despite ROS and NETs having been inhibited.

Recent doubt has been cast on the specificity of 3PO for its reported target PFKFB3 [31, 32]. When we repeated our experiments with a highly specific PFKFB3 inhibitor we observed no effect on ROS or NET production, leading us to consider that 3PO may have an off-target, anti-inflammatory effect in human neutrophils. Our NMR metabolomics analysis confirmed an increase in lactate in response to 3PO, which aligns with recent reports [31]. We also observed a significant decrease in glutathione, which is an important metabolite in redox homeostasis. It is essential for defending cells against oxidative damage, including from ROS such as hydrogen peroxide and superoxide produced during the neutrophil respiratory burst. It acts directly as an antioxidant and as a substrate for glutathione peroxidases (GPXs), which break down peroxides and prevent oxidative damage to biomolecules [52]. The decrease in glutathione observed in our experiments is likely in direct response to the decrease in ROS production in the presence of 3PO. The increase in mitochondrial metabolites L-glutamine and phenylalanine, along with the predicted increase in amino acid-recycling, may represent a metabolic switch adopted by neutrophils to compensate for the inhibition of glycolysis. L-glutamine is a significant energy source for cells of the immune system, where it is converted to glutamate and then into alpha-ketoglutarate, before entering the TCA cycle to produce ATP [53]. In rat neutrophils, glutamine enhances ROS production and regulates NADPH oxidase activity [54]. It is interesting therefore, that the levels of glutamine increase in response to 3PO and yet ROS production is completely inhibited. Phenylalanine and its metabolite tyrosine are degraded to fumarate and acetoacetate, which are also intermediates in the TCA cycle, thus contributing to the production of ATP in the absence of glycolysis. Whilst we did not detect TCA cycle activity in PMA-activated neutrophils in our Seahorse experiments, it may be that some TCA enzymes are functional in neutrophils and may become activated in conditions of glycolysis inhibition. Further experiments should confirm whether intermediary enzymes of the TCA cycle can become active under stress conditions in neutrophils. Metabolic pathway enrichment analysis also predicted changes in a number of metabolic pathways involving membrane phospholipids, possibly reflecting the changes in the proportion of early/late apoptotic neutrophils observed in our study.

One of the limitations of our study was that the participants with RA were receiving one or more disease-modifying anti-rheumatic drugs (DMARDs e.g. methotrexate and hydroxychloroquine), and this may account for some of the heterogeneity in the results for the RA group. The treatment groups and exclusion criteria for HC and RA samples also lead to modest sample size that limits the effectiveness of univariate analysis methods for metabolomics datasets. Data filtering using CRS to limit analysis to annotated metabolites within the data (53 metabolites) did not wholly alleviate this issue and therefore it is possible that more metabolite changes could be discerned with a larger sample size. Several DMARDs have known effects on neutrophil functions, including decreasing ROS production and degranulation [9]. We recorded wider experimental variation in the RA samples, which could be explained by the effect of DMARDs, differences in donor disease activity (DAS28) or other clinical factors such as inflammation (e.g. CRP). We were also not able to conclusively determine the molecular target of 3PO in human neutrophils.

In summary, we have shown in this study that neutrophils are metabolically adapted to maintain ROS production in low glucose environments such as sites of inflammation, and that NET production is preserved in RA neutrophils in low glucose environments. We also identified the inhibitor compound 3PO as a critical regulator of pathogenic and tissue damaging inflammatory responses by human neutrophils. Further experiments will be required to fully determine whether this is via direct inhibition of PFKFB3, or via an off-target effect, which may divert energy production away from glycolysis and towards mitochondrial metabolism. Whilst 3PO partially inhibited cytotoxic bacterial killing by neutrophils we propose that, at lower and optimised concentrations, 3PO could be considered a potential therapeutic for the treatment of immune-mediated inflammatory diseases such as RA with a strong neutrophil-driven pathology.

## 6. ACKNOWLEDGEMENTS

We would like to thank the rheumatology nurses and consultants at Liverpool University Hospital NHS Foundation Trust for their assistance in recruiting patients for this study. We thank Dr Amy Chadwick for use of the Seahorse XFe96 analyser. We would like to thank Prof Steven Edwards for constructive and critical discussions throughout this project.

## 7. ETHICAL APPROVAL

The study was approved by the University of Liverpool Central University Research Ethics Committee C and NRES Committee North West (Greater Manchester West, Manchester, UK) for respectively the isolation of neutrophils from HC and RA blood. All participants gave written, informed consent in accordance with the declaration of Helsinki. All patients fulfilled the ACR 2010 criteria for the diagnosis of RA and were Biologics naïve. People with RA (60% female) were recruited from Liverpool University Hospital Foundation Trust in Liverpool, UK and had clinician-diagnosed, active disease at the time of participation. Healthy controls (54% female) were recruited from volunteers and colleagues at the University of Liverpool. Participants were aged 18-75 years, and did not have an active infection.

## 8. AUTHOR CONTRIBUTIONS

MFA designed the experiments, carried out the experiments, analysed the data and wrote the manuscript. GA carried out the experiments, analysed the data and revised the manuscript. AN, CL, MM, AC, AS carried out the experiments and revised the manuscript. MMP designed the experiments, analysed the data and wrote the manuscript. HLW designed the research, performed the experiments, analysed the data, and wrote the manuscript.

## 9. FUNDING

MFA was funded by a Versus Arthritis and Masonic Charitable Fund PhD scholarship (Grant No. 22193). GA was funded by a University of Liverpool CIMA MRes scholarship and a Dunhill Medical Trust and University of Liverpool PhD Scholarship. AN was funded by a Connect Immune Research and The Lorna & Yuti Chernajovsky Biomedical Research Foundation Grant (Grant No. 22925). HLW was funded by a Versus Arthritis Career Development Fellowship (Grant No. 21430). The equipment and software licences used in the Shared Research Facility for NMR metabolomics were funded by the MRC (Grant No. MR/M009114/1).

## 10. DATA AVAILABILITY

All data presented in this manuscript are available upon reasonable request to the corresponding author.

## 11. CONFLICT OF INTEREST

The authors declare no conflicts of interest. All grant funding for the research has been declared. The authors have no financial relationships with any organizations that might have an interest in the submitted work in the previous three years. No other relationships or activities that could appear to have influenced the submitted work.

## Notes

### Competing Interest Statement

The authors have declared no competing interest.

### Author Declarations

The study was approved by the University of Liverpool Central University Research Ethics Committee C and NRES Committee North West (Greater Manchester West, Manchester, UK) for respectively the isolation of neutrophils from HC and RA blood. All participants gave written, informed consent in accordance with the declaration of Helsinki.

### Summary of Updates

Additional experimental analysis Figure 4E, Figure 6 and Figure 7. Figure 8 was previously figure 7. The discussion and conclusions have been revised in light of additional data analysis.

